# Lesion location changes the association between brain excitability and motor skill acquisition post-stroke

**DOI:** 10.1101/2024.07.30.24311146

**Authors:** Bernat De Las Heras, Lynden Rodrigues, Jacopo Cristini, Kevin Moncion, Numa Dancause, Alexander Thiel, Jodi D. Edwards, Janice J. Eng, Ada Tang, Marc Roig

## Abstract

**Background:** The capacity to reacquire motor skills lost after a stroke is crucial to promote upper-limb motor recovery but the impact of lesion location on motor skill acquisition and the underlying neurophysiological mechanisms remain uncertain.

**Methods:** We used transcranial magnetic stimulation to investigate associations between excitatory and inhibitory cortico-spinal excitability measures and the capacity to acquire a novel motor skill with the most affected hand in 103 individuals with cortical (n=34) or subcortical (n=69) lesions.

**Results:** Both groups showed similar motor skill acquisition, but subcortical lesions exhibited more impairment in the most affected hand and lower excitability in the ipsilesional hemisphere. In cortical lesions, motor skill acquisition was associated with lower motor thresholds (β=-0.25, 95% CI [−0.47,-0.03]; p=0.024) and higher intracortical inhibition (β=-3.93, 95% CI [−6.89,-0.98]; p=0.011) in the ipsilesional hemisphere. In contrast, in subcortical lesions motor skill acquisition was associated with smaller motor evoked potentials (β=-4.46, 95% CI [−8.54,-0.38]; p=0.033), less intracortical inhibition (β=3.45, 95% CI [0.34,6.56]; p=0.030) and higher facilitation (β=1.34,95% CI [0.15,2.54]; p= 0.028) ipsilesionally. Sensitivity analyses revealed that associations with intracortical inhibition and facilitation in the subcortical group were driven by lesions affecting the corticospinal tract. No associations were found in the contralesional hemisphere.

**Conclusions:** Reinforcing the existence of lesion-specific neurophysiological patterns, individuals with cortical and subcortical lesions show divergent associations between cortico-spinal excitability and motor skill acquisition. The use of cortico-spinal excitability as a biomarker to predict upper-limb recovery post-stroke or guide motor recovery interventions such as non-invasive brain stimulation should consider lesion location.

## Introduction

Stroke is a leading cause of disability worldwide, with up to 80% of patients experiencing upper-limb motor deficits in acute stages of recovery, and only around half achieving full upper-limb recovery at six months after the cerebrovascular accident.^1^ Upper-limb motor impairments can lead to difficulties in the execution of motor skills that are essential for activities of daily living such as reaching or holding objects and thus reduce functional independence and health-related quality of life.^2^

Although motor learning, defined as the capacity to acquire and retain motor skills,^3^ is not the same as motor recovery, similar neurobiological mechanisms underlie both processes. Studies demonstrate that comparable changes in neuronal excitability occur in both motor learning and post-stroke motor recovery.^4^ These studies support the view that the mechanisms underlying motor learning provide a substrate for motor recovery and thus guide motor rehabilitation post-stroke.^3^

Shared mechanisms supporting motor learning and stroke recovery have been well characterized in animal studies but are less understood in humans.^5^ This is in part because while stroke lesions induced on animals can be rigorously localized and controlled, stroke lesions in humans have greater neuroanatomical variability, which often results in a broader range of clinical outcomes. Reducing this heterogeneity by controlling for relevant lesion-related characteristics could improve our understanding of mechanisms underlying upper-limb recovery post-stroke.^6^

Lesion location plays a fundamental role in determining both initial impairment and motor recovery after stroke.^7^ The ability to regain upper-limb motor function is strongly dependent on the functional integrity of the corticospinal tract (CST).^8^ Compared with lesions affecting cortical areas, subcortical lesions affecting the CST tend to result in larger impairments and poorer motor recovery.^7^ Individuals with subcortical lesions also tend to show a more widespread brain activation during upper-limb paretic movements,^9^ with more damage in the CST leading to an increased recruitment of secondary motor networks.^10^

Given its capacity to assess the functional integrity of the CST, transcranial magnetic stimulation (TMS) has been extensively used in stroke trials.^11^ Cortico-spinal excitability (CSE) measures obtained with single and paired pulse TMS protocols have been used to characterize motor impairment and upper-limb recovery post-stroke^8^ as well as to assess intracortical facilitation and inhibition mechanisms, providing insights into excitatory (glutamate) and inhibitory (g-aminobutyric acid -GABA-) neurotransmitters^12^ involved in motor learning and motor recovery (**Fig. 1**).^13^

**Fig. 1.**
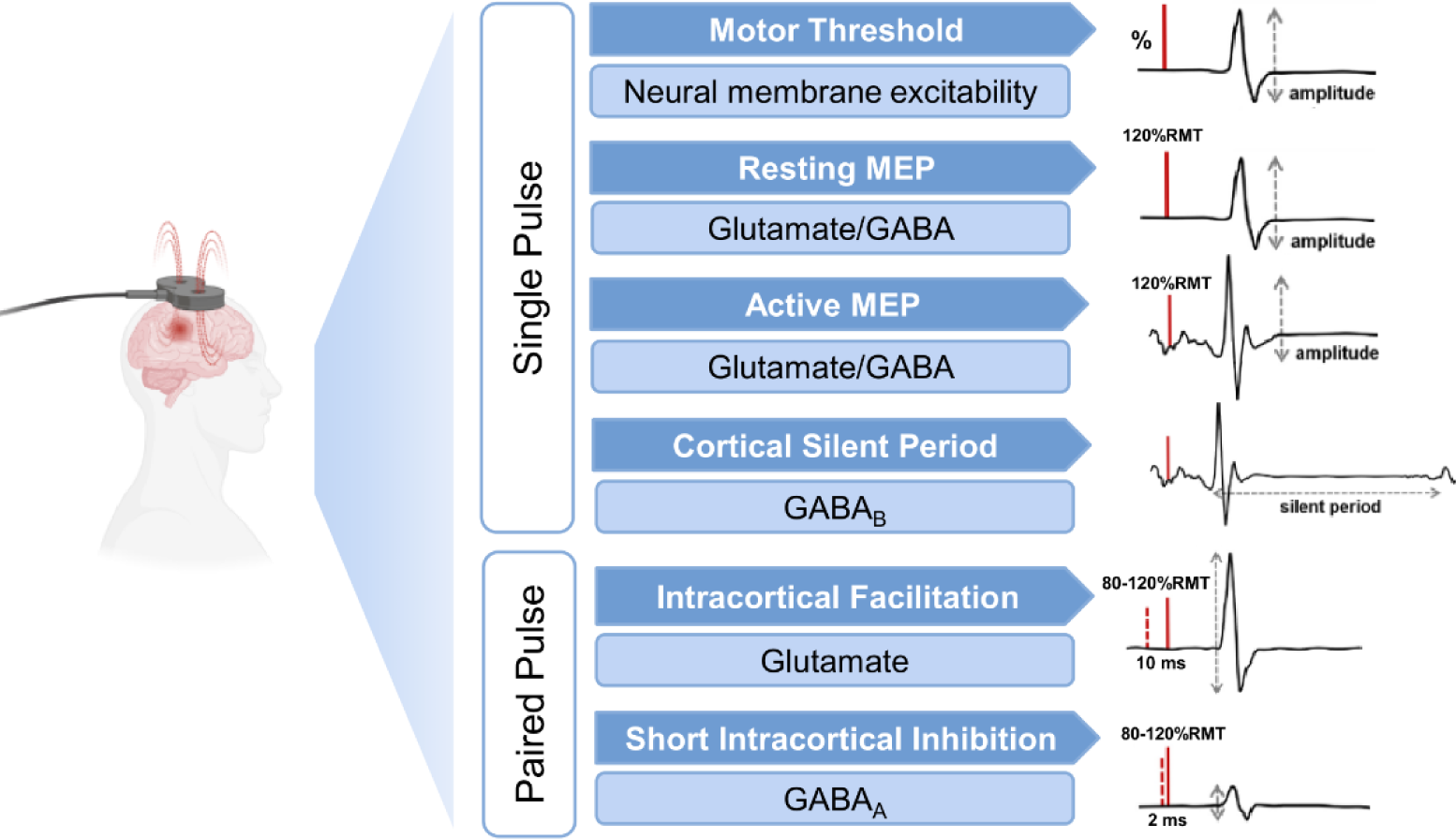
Single and paired pulse transcranial magnetic stimulation protocols used to assess different corticospinal excitability (CSE) measures and their putative underlying mechanisms. Resting Motor Threshold (RMT) is shown as a percentage of the stimulator output capacity, reflecting neural membrane excitability, with lower RMT indicating higher CSE. Motor evoked potential (MEP) amplitude measures excitability of cortical and spinal projections influenced by excitatory (glutamate) and inhibitory (GABA) circuits, with larger MEP amplitudes indicating higher CSE. Cortical Silent Period (CSP) reflects GABA_B_ receptor-mediated inhibition, with longer CSP indicating greater inhibition. Intracortical Facilitation (ICF) measures facilitation mediated by NMDA receptors, and SICI (Short Intracortical Inhibition) assesses inhibition mediated by GABA_A_ receptors. Larger ICF values indicate greater facilitation, while larger SICI values indicate greater inhibition. GABA, gamma-aminobutyric acid; ms, milliseconds; NMDA, N-methyl-D-aspartate.

Lesion location has been shown to influence CSE, with lesions involving cortical and subcortical brain structures differing in excitability patterns.^14^ Lesion location has also been shown to alter the association between CSE and upper-limb motor impairment.^15^ Despite the importance of motor learning for upper-limb recovery post-stroke,^16^ whether lesion location influences the association between CSE and motor skill acquisition has not yet been investigated. We used TMS to study the influence of lesion location on the association between CSE and upper-limb motor skill acquisition in individuals with cortical and subcortical lesions at different stages of the stroke recovery continuum.

## Methods

### Experimental Design

This cross-sectional study includes baseline data from individuals participating in two registered randomized control trials (NCT03614585, NCT05076747) and adheres to the relevant STROBE checklist.^17^ In the first session, in addition to descriptive measures, stroke severity, cognitive function, upper-limb impairment and function were assessed with the National Institutes of Health Stroke Scale (NIHSS), the Montreal Cognitive Assessment (MoCA), the arm and hand dimensions of the Chedoke-McMaster Stroke Assessment (CMSA), and the Box and Block Test (BBT), respectively. Motor skill acquisition was assessed with a time-on-target motor task using the hand contralateral to the lesioned hemisphere. Two days after the first experimental session TMS was applied over the primary motor cortex (M1) of the ipsilesional and contralesional hemispheres to obtain different CSE measures. An ethics review board approved the project and participants provided written consent prior to participation according to the Declaration of Helsinki.

### Participants

We included individuals with first-ever ischemic or hemorrhagic stroke, from early subacute to chronic stages (7 days to 5 years post-stroke) of recovery, with no upper-limb musculoskeletal or neurological conditions other than stroke-related motor deficits, and no TMS contraindications.^18^ Lesion location was determined from CT/MRI scans obtained ≤2 days after the stroke event, and confirmed by a clinical radiologist. The subcortical group included individuals with lesions involving only subcortical regions. To conduct exploratory sensitivity analyses, this group was further divided between participants with lesions affecting the CST (e.g., internal capsule, corona radiata) and participants with subcortical lesions outside this pathway (e.g., basal ganglia, thalamus). The cortical group included individuals with lesions involving cortical regions (e.g., frontal, parietal, occipital) as well as those with combined cortical/subcortical infarcts.^14,15^ Individuals with lesions located in the cerebellum or brainstem were excluded.

### Maximal Voluntary Contraction

Handgrip maximal voluntary contraction (MVC) force was assessed for both hands using a custom LabView script. Patients, seated with a handgrip force response pad, viewed a slider on a 27-inch screen that provided visual feedback on force produced. They performed three 3-second MVCs with 30-second pauses. The highest MVC was recorded.^19^

### Time-on-target Motor Task

To assess motor skill acquisition, participants used a time-on-target motor task requiring fine hand-grasping force modulation (**Fig. 2**). with the same handgrip force response pad used for the MVC assessment with the hand contralateral to the lesioned hemisphere.^19^ Participants were instructed to increase or release grip force to maintain the cursor within targets for as long as possible while keeping the grip position as steady as possible.

**Fig. 2.**
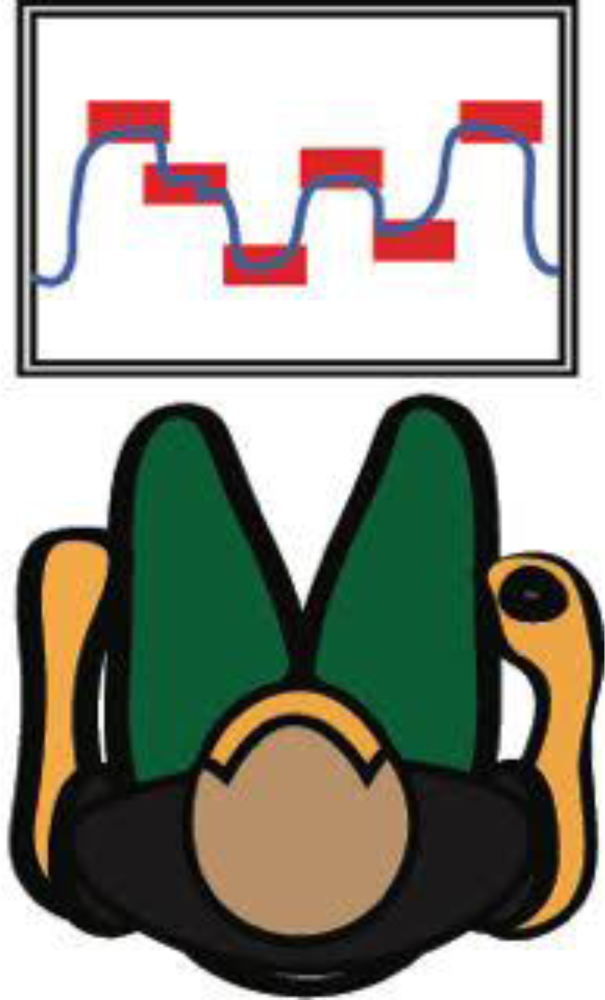
Time on target motor task used to assess motor skill acquisition. The blue cursor crossed a 27-inch computer screen from left to right at a constant speed of 8 seconds/screen. Participants needed to apply or release grip force to adjust the blue cursor up or down in order reach the 12 red targets displayed horizontally at different heights. The force required to reach the highest target was ∼20% of MVC. The goal was to keep the cursor within the targets as much time as possible.

To minimize skill level differences at baseline, before practice, participants were allowed to perform familiarization trials until they scored ≥30 at least three times. Practice consisted of 4 blocks of 20 trials each with 1-2 minutes of rest between blocks.^19^ Excluding familiarization, total practice time was 21.6 minutes. The score, which was visually presented at the end of each trial, was calculated as time on target divided by total time of each trial multiplied by 100. The difference in mean scores from the first block to the best block of practice was used as measure of motor skill acquisition.^19^

### Transcranial Magnetic Stimulation

Using neuronavigation (Brainsight, Rogue Research Inc., Montreal, QC, Canada), we first co-registered the patients’ heads to a standard MRI template to identify and mark the optimal coil position (“hot-spot”) of M1 for eliciting motor-evoked potentials (MEP).^20^ TMS was applied through a 70-mm coil with a Bistim^2^ stimulator (Magstim, Whitland, Wales, UK), oriented posteriorly at a 45⁰ angle relative to the midsagittal line targeting the M1 representational area of the first dorsal interosseous muscle in both the ipsilesional and contralesional hemispheres.^21^

Electromyographic activity was recorded via two surface electrodes placed over the first dorsal interosseous ∼1 cm apart. Data was acquired was acquired at 2000 Hz through a CED Micro1401-4 unit, controlled by Signal software (CED, Cambridge, UK), with a gain of 300 and filtered using 10 Hz high-pass and 500 Hz low-pass filters. Different CSE measures were obtained via single and paired-pulse TMS protocols (**Fig. 1**).

### Corticospinal Excitability

#### Single-pulse protocols

*<u>Resting motor threshold (RMT)</u>:* defined as the minimum stimulation intensity required to elicit MEPs of >0.05 mV in at least 10 of 20 trials.^21^ *<u>MEP amplitude</u>:* assessed both at rest and during an active muscle contraction sustained at 10% of the MVC. To assess active excitability, the LabView script used to measure MVC provided visual feedback while participants were asked to maintain the muscle contraction at the 10% MVC level. MEP amplitude was quantified by averaging the peak-to-peak amplitude from 60 stimulations (30 resting and 30 active) delivered 5 seconds apart at 120% RMT.^22^ *<u>Cortical silent period (CSP)</u>*: the CSP is a period of electrical silence in the electromyographic (EMG) activity following an MEP during isotonic muscle contraction.^21^ CSP was extracted from 30 stimulations at 120% RMT during an active contraction. The EMG baseline signal amplitude 200 ms before stimulation was measured. The end of the MEP and the recovery of the voluntary EMG activity (i.e., two standard deviations above the mean baseline signal amplitude) marked the beginning and the end of the CSP, respectively.^19^

#### Paired-pulse protocols

*<u>Intracortical facilitation (ICF) and short intracortical inhibition (SICI):</u>* measured using a conditioning pulse of 80% RMT followed by a suprathreshold pulse (120% RMT) delivered at rest after 10-12 (ICF) and 2-2.5ms (SICI), respectively. The amplitude of the MEP elicited by the second pulse was normalized to the unconditioned resting MEP amplitudes at 120% RMT to estimate facilitation and inhibition.^19^ A total of 60 paired-pulses (30 for ICF and 30 for SICI) were delivered with an interstimulus interval of 5 seconds.^21^

### Statistical Analysis

Data were visually inspected with histograms and normal quantile plots and the Shapiro-Wilk test was used to confirm normality of distribution for each variable. Differences between cortical and subcortical groups in demographic and clinical variables were investigated using t-tests and Wilcoxon tests for continuous variables. Chi-square tests (X^2^) were used to compare groups in categorical variables.

Repeated linear mixed models (LMM) were used to assess differences between groups in motor skill acquisition. Scores of practice blocks (Block 1 to 4) were the dependent variable, and block, group, and their interaction, were treated as fixed effects, and participants as a random effect. Stroke severity (NIHSS), age, time since stroke (days), and handgrip MVC were entered as covariates. Auto Regressive order 1 (AR1) was set as the repeated covariance structure.

LMMs were used to assess differences between groups for CSE. In this case, each specific CSE measure was the dependent variable and group was the fixed effect. Stroke severity (NIHSS), age, and time since stroke (days) were the covariates entered in the models. Handgrip MVC was not included as covariate to reduce redundancy. All LMMs treated participants as a random effect.

Multivariate linear regression models were employed to investigate associations between motor skill acquisition and CSE for cortical and subcortical groups, including the same covariates as in the motor skill acquisition LMMs. Separate models analyzed CSE from ipsilesional and contralesional hemispheres for each group. Multicollinearity between covariates was assessed with the variance inflation factor (VIF), with a threshold of >5 indicating excessive multicollinearity. Linear model assumptions were checked for residual normality, and influential observations were identified using leverage plots and Cook’s Distance (score >1).

Exploratory sensitivity analyses compared individuals with cortical lesions to those with subcortical lesions affecting only CST-associated regions or regions without direct CST involvement, examining their impact on motor skill acquisition, CSE, and their association. The main outputs of these (three-group) analyses, are summarized here and detailed in supplementary files. All analyses were performed with two-tailed test using JMP (version 17) from SAS. The alpha level was set at <0.05.

## Results

### Sample characteristics

After removing from the analysis 13 individuals with lesions affecting the cerebellum and/or brainstem, a total of 103 participants were classified based on lesion location as cortical (n=34) or subcortical (n=69) (**Table 1**). Within the subcortical group, 26 participants had lesions affecting the CST and 43 participants lesions not affecting this structure. Time since stroke (days) was longer in the subcortical group, although this difference became non-significant when groups were stratified as subacute (<6 months) and chronic (>6 months) categories. Non-statistically significant differences were observed between cortical and subcortical groups for any other demographic variables. However, in individuals with subcortical lesions, measures of upper-limb motor impairment (CMSA_arm,_ CMSA_hand_), hand strength (MVC_affected_) and function (BBT_affected_) of the most affected hand exhibited larger deficits (**Table 1)**.

**Table 1.** Demographic and clinical data for cortical and subcortical groups.

|  | **Cortical** | **Subcortical** | **p value** |
| --- | --- | --- | --- |
| **N** | 34 | 69 |  |
| **Age (y)** | 64.79 (11.61) | 65.27 (9.51) | **0.823** |
| **Sex (F/M)** | 7/27 | 25/44 | **0.107** |
| **Subacute/Chronic** | 28/6 | 41/28 | **0.020\*** |
| **Days since stroke** | 135.94 (199.85) | 315.13 (429.86) | **0.025\*** |
| **Subacute** | 63.64 (22.85) | 65.29 (23.11) | **0.292** |
| **Chronic** | 473.33 (308.08) | 680.96 (480.64) | **0.321** |
| **Type of stroke (I/H)** | 33/1 | 60/9 | **0.103** |
| **Lesion location** | Frontal = 29.6%<br>Parietal= 14.7%<br>Occipital= 5.8%<br>Multiple cortical regions= 14.7%<br>Cortico-Subcortical=35.2% | Corona radiata= 17.4%<br>Basal ganglia= 21.8%<br>Internal capsule=18.7%<br>Thalamus=10.9%<br>Multiple subcortical regions= 31.2 % |  |
| **NIHSS** | 1.79 (1.73) | 1.97 (2.09) | **0.671** |
| **MoCA** | 23.75 (4.95) | 23.56 (4.98) | **0.857** |
| **CMSA<sub>arm</sub>** | 6.54 (1.09) | 5.41 (1.80) | **0.0004\*** |
| **CMSA<sub>hand</sub>** | 6.42 (0.87) | 5.32 (1.88) | **0.0061\*** |
| **MVC<sub>affected</sub>** | 0.79 (0.25) | 0.63 (0.31) | **0.0070\*** |
| **MVC<sub>unaffected</sub>** | 0.86 (0.21) | 0.83 (0.29) | **0.641** |
| **BBT<sub>affected</sub>** | 49.57 (11.44) | 41.40 (15.33) | **0.0100\*** |
| **BBT<sub>unaffected</sub>** | 52.12 (7.74) | 49.72 (9.67) | **0.221** |

BBT, Box and Block Test; CMSA, Chedoke-McMaster Stroke Assessment; M, Male; MoCA, Montreal Cognitive Assessment; MVC, Maximal voluntary contraction; NIHSS, National Institutes of Health Stroke Scale; F, Female. Values are presented as means and standard deviations (SD). \* $p<0.05$ .

The exploratory sensitivity analyses indicated no significant differences among the three lesion groups (cortical, subcortical with CST affected, and subcortical with CST unaffected) in age (F_(2,100)_=0.09; p=0.906), sex (X^2^_(2,103)_=3.262; p=0.196), time since stroke (F_(2,100)=_2.75; p=0.063), stroke severity (NIHSS) (F_(2,100)_=0.20; p=0.819), and cognitive status (MoCA) (F_(2,100)_=3.05; p=0.052). However, participants with CST subcortical lesions showed significantly worse CMSA_arm_ (F_(2,94)_=9.50; p=0.0002), CMSA_hand_ (F_(2,95)_=7.89; p=0.0007), MVC_affected_ (F_(2,93)_=5.42; p=0.0060) and BBT_affected_ (F_(2,96)_=4.06; p=0.0203) (**Supplementary Table 1)**.

### Motor skill acquisition

There were no significant differences between the cortical and subcortical groups in motor skill performance at baseline (Block 1) (t=0.40; p=0.999). Similarly, motor skill acquisition was not significantly different between groups (F_(3,169.9)_=1.27, p=0.287), with the cortical group showing an improvement of 8.26 (1.25) score points and the subcortical group 5.91 (0.90) score points during motor practice **(Fig. 3)**. Differences in motor skill acquisition between cortical and subcortical groups remained insignificant even when LMMs were not adjusted with covariates (F_(3,188.7)_=1.09, p=0.353)(**Supplementary Tables 2.1-2.2**). Similarly, in the exploratory sensitivity analyses, both adjusted (F_(6,193.3)_=0.70, p=0.652) an unadjusted (F_(6,212.7)_=0.82, p=0.552) LMMs also revealed no differences among the three lesion groups in motor skill acquisition (**Supplementary Tables 2.3-2.4**).

**Fig. 3.**
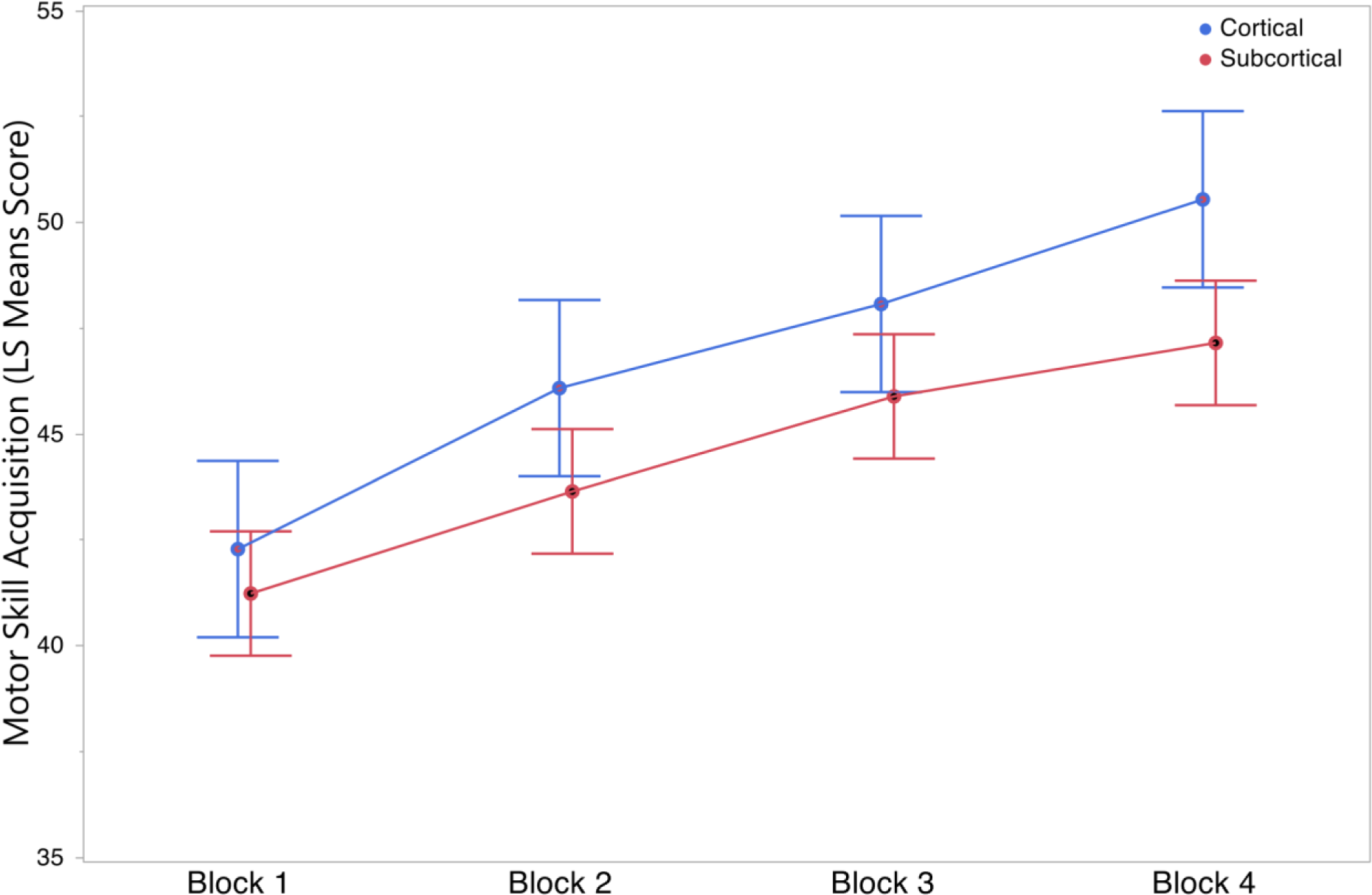
Group scores in motor skill acquisition. Data is presented as least squares means estimates with standard errors of the mean. LMMs were adjusted for stroke severity, age, handgrip maximal voluntary contraction and time since stroke.

### Cortico-spinal excitability

Cortico-spinal excitability (CSE) measures were obtained from all participants except 5, whose MEPs could not be elicited on neither hemisphere. These participants were excluded from CSE analyses but included in the motor skill acquisition analysis. For 10 participants with no response in the ipsilesional hemisphere, only contralesional hemisphere data were used. Participants with subcortical lesions tended to have higher RMT and smaller resting and active MEP amplitude. However, only active MEP significantly differed between cortical and subcortical groups (**Table 2**). Differences in ipsilesional CSE measures of inhibition (CSP and SICI) and facilitation (ICF), as well as contralesional CSE measures, were not statistically significant. Details on the LMMs comparing CSE measures between cortical and subcortical groups are provided in **Supplementary Table 3**.

**Table 2.** Cortico-spinal excitability (CSE) measures in ipsilesional and contralesional hemispheres.

|  | **Cortical** | **Subcortical** | **p value** |
| --- | --- | --- | --- |
| **Ipsilesional** |  |  |  |
| **RMT (%)** | 48.84 (2.19) | 54.16 (1.59) | **0.056** |
| **Resting MEP (mV)** | 0.51 (0.07) | 0.38 (0.05) | **0.126** |
| **Active MEP (mV)** | 1.74 (0.15) | 1.33 (0.11) | **0.031\*** |
| **CSP (ms)** | 0.19 (0.01) | 0.19 (0.01) | **0.982** |
| **ICF (mV)** | 1.71 (0.20) | 1.91 (0.15) | **0.439** |
| **SICI (mV)** | 0.93 (0.11) | 0.74 (0.08) | **0.195** |
| **Contralesional** |  |  |  |
| **RMT (%)** | 51.74 (1.91) | 51.38 (1.32) | **0.878** |
| **Resting MEP (mV)** | 0.47 (0.04) | 0.41 (0.03) | **0.303** |
| **Active MEP (mV)** | 1.71 (0.14) | 1.76 (0.10) | **0.772** |
| **CSP (ms)** | 0.18 (0.01) | 0.17 (0.01) | **0.255** |
| **ICF (mV)** | 1.73 (0.23) | 1.90 (0.16) | **0.569** |
| **SICI (mV)** | 0.54 (0.16) | 0.83 (0.11) | **0.141** |

CSP, cortical silent period; ICF, intracortical facilitation; MEP, motor evoked potential; RMT, resting motor threshold; SICI short-intracortical inhibition. Values are means estimates and standard errors of the mean (SEM). LMMs were adjusted for stroke severity, age and time since stroke \* $p < 0.05$ .

The results of the exploratory sensitivity analyses revealed that participants with lesions involving the CST tended to show less excitability in the ipsilesional hemisphere, as indicated by significantly smaller resting (F_(2,82)_=4.92, p=0.0091) and active (F_(2,82)_=8.29, p=0.0005) MEP amplitudes (**Supplementary Table 4**). RMT was also higher in this group, although differences did not reach statistical significance. No significant differences among the three groups were found in the other ipsilesional or contralesional CSE measures. Details on the LMMs comparing CSE among the three lesion groups are provided in **Supplementary Table 5**.

### Associations between CSE and skill acquisition

Divergent associations between several CSE measures and motor skill acquisition were observed in cortical and subcortical groups in the ipsilesional hemisphere (**Fig. 4**). In individuals with cortical lesions, motor skill acquisition was associated with lower RMT (β=-0.25, 95% CI [−0.47, −0.03]; p=0.024) and increased intracortical inhibition (β=-3.93, 95% CI [−6.89, −0.98]; p=0.011) in the ipsilesional hemisphere. In contrast, in individuals with subcortical lesions motor skill acquisition was associated with smaller resting MEP amplitude (β=-4.46, 95% CI [−8.54, - 0.38]; p=0.033), increased intracortical facilitation (β=1.34, 95% CI [0.15,2.54]; p= 0.028), and reduced intracortical inhibition (β=3.45, 95% CI [0.34,6.56]; p=0.030) in the ipsilesional hemisphere. In the contralesional hemisphere neither the cortical or subcortical group exhibited any significant association between any CSE measure and motor skill acquisition (**Fig. 5)**.

**Fig. 4.**
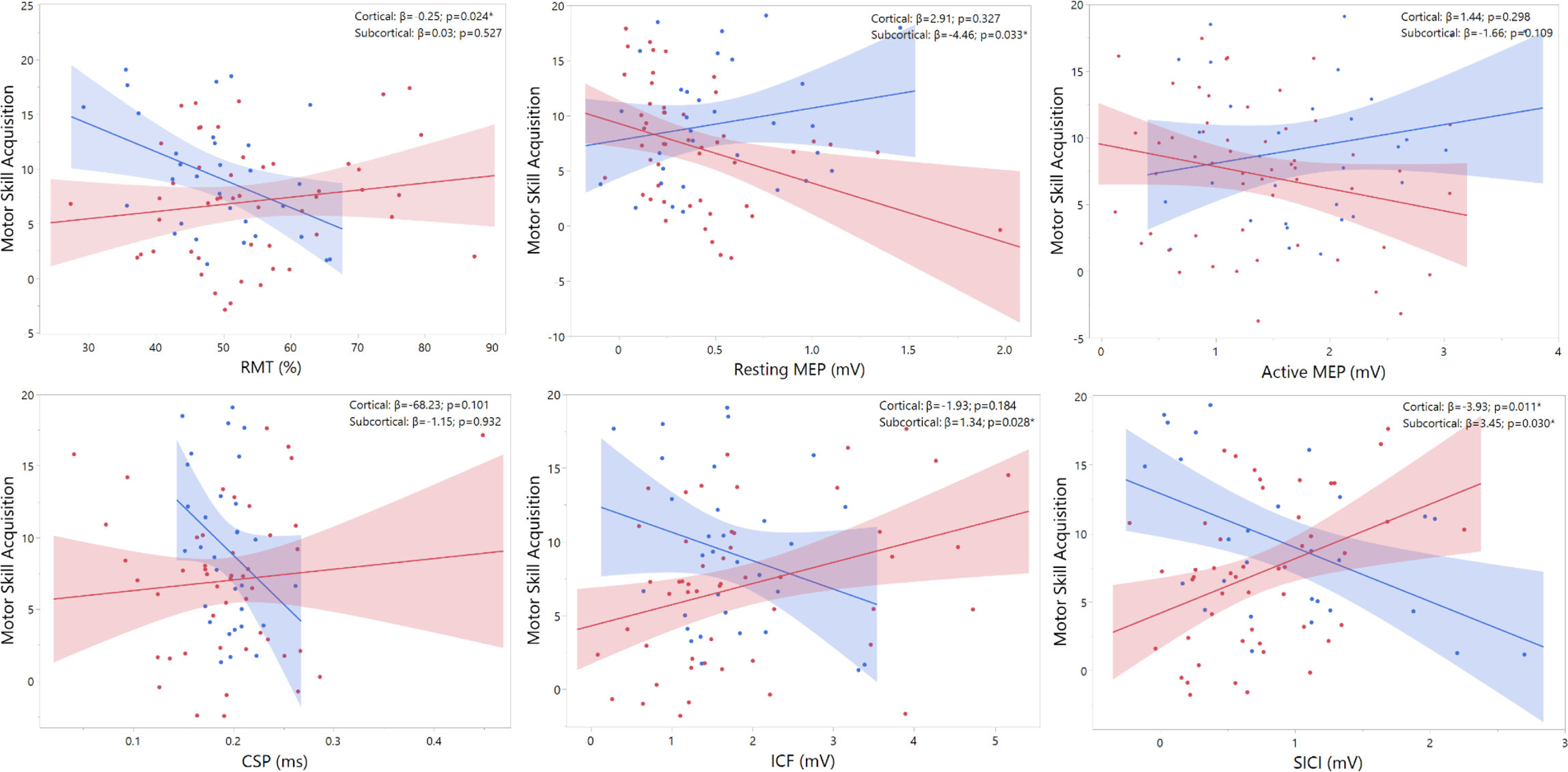
Partial regression plots showing the associations between each CSE measure in the ipsilesional hemisphere and motor skill acquisition in cortical and subcortical lesions. Cortical and subcortical groups are depicted in blue and red, respectively. CSP, cortical silent period; ICF, intracortical facilitation; MEP, motor evoked potential; RMT, resting motor threshold; SICI, short-intracortical inhibition. Note that in ICF and SICI conditioned MEP amplitude is normalized to the unconditioned resting MEP amplitude and that larger ICF and SICI values represent higher facilitation and lower inhibition, respectively. * p<0.05.

**Fig. 5.**
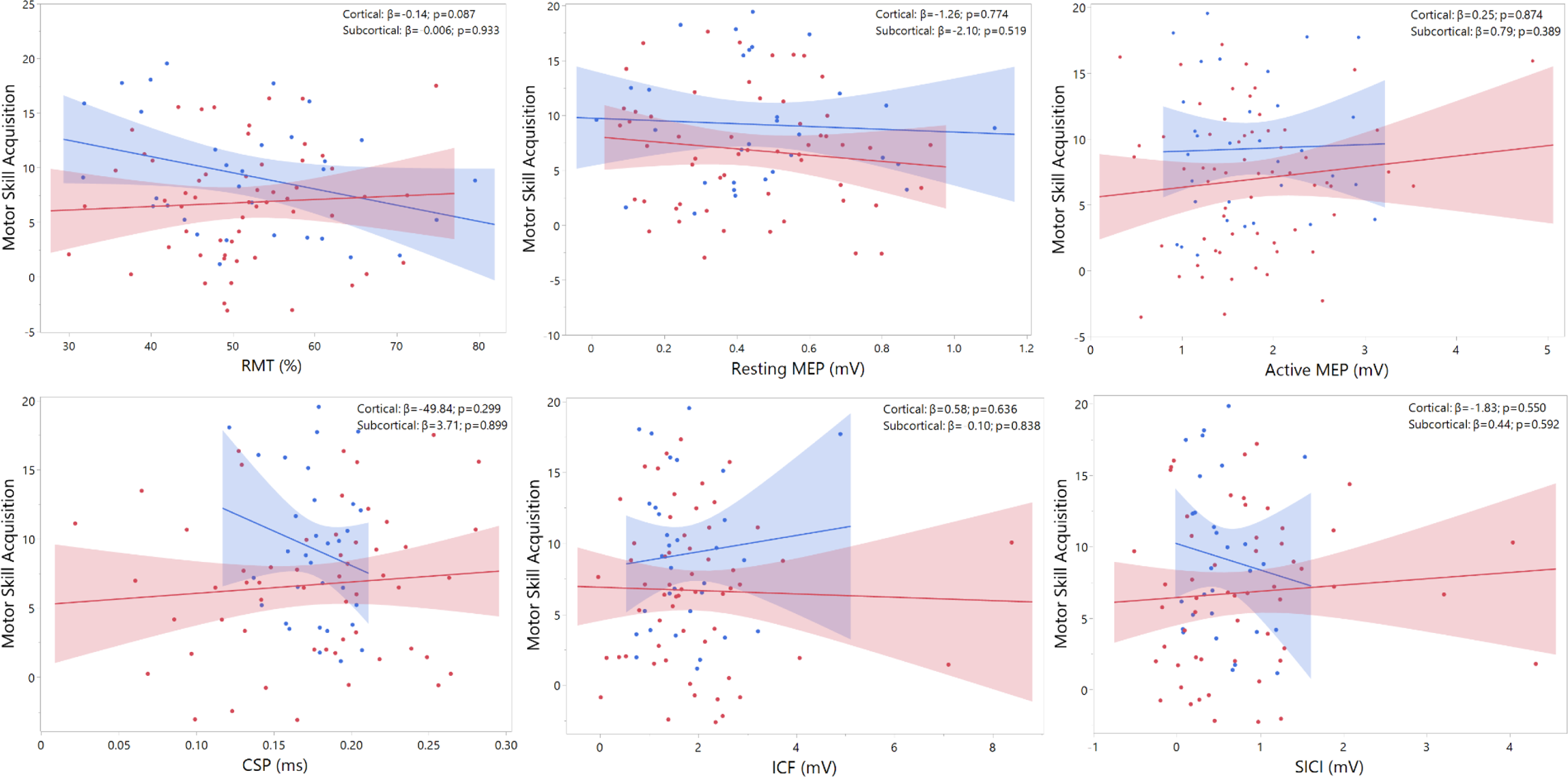
Partial regression plots showing the associations between each CSE measure in the contralesional hemisphere and motor skill acquisition in cortical and subcortical lesions. Cortical and subcortical groups are depicted in blue and red, respectively. CSP, cortical silent period; ICF, intracortical facilitation; MEP, motor evoked potential; RMT, resting motor threshold; SICI, short-intracortical inhibition. Note that in ICF and SICI conditioned MEP amplitude is normalized to the unconditioned resting MEP amplitude and that larger ICF and SICI values represent higher facilitation and lower inhibition, respectively. * p<0.05.

The exploratory sensitivity analyses revealed that significant associations between CSE and motor skill acquisition within the subcortical group were driven by observations from individuals with CST lesions. Associations with SICI (β=10.31, 95% CI [2.07,18.54]; p=0.019) and ICF (β=2.64, 95% CI [0.34,4.94]; p=0.027) remained significant when only the data of the subcortical group with CST lesions were analyzed. No significant associations were found for any CSE measure on the contralesional hemisphere. Details of all regression models are provided in **Supplementary Tables 6-9**.

## Discussion

The CST is a collection of axons stemming from multiple territories in the sensorimotor cortex, primarily M1, but also from premotor areas and supplementary motor areas, descending in a funnel-like manner to converge subcortically into the corona radiata, internal capsule and peduncles.^23^ Damage to the CST is the common denominator for post-stroke hemiparesis and can lead to significant deficits in upper-limb function, with studies identifying its degree of lesion as the best predictor of precision grip function.^2^ Taken together, our findings support the view that lesions in subcortical brain areas where descending motor pathways are densely compacted are more likely to produce upper-limb impairments than more superficial cortical lesions and that subcortical lesions affecting the CST tend to augment these impairments.^7^

We assessed motor skill acquisition using a handgrip task that mimics essential activities of daily living such as grasping and manipulating objects.^19^ The performance of these tasks, which are correlated with upper-limb function, is thought to rely on the integrity of the CST.^2^ Considering the influence that descending corticospinal projections and subcortical brain regions have on motor skill acquisition,^24^ in upper-limb reaching and hand manipulation tasks, one would expect worse motor skill acquisition in the subcortical group, especially if the lesion affects the CST.^25^ However, while performance tended to be lower in individuals with subcortical lesions, acquisition rates did not differ between groups. This result aligns well with previous studies suggesting a relatively well-preserved motor skill learning capacity post-stroke.^26,27^

Differences between groups in CSE emerged only in the ipsilesional hemisphere, with the subcortical group showing smaller MEP amplitudes assessed during the performance of an active contraction (**Table 1**). It is possible that this reduced CSE becomes only apparent during muscle contraction due to spinal facilitation, which increases MEP amplitudes when active force is produced.^21^ Despite using significantly higher TMS intensities, the subcortical group also showed a tendency to produce higher RMTs and smaller resting MEP amplitudes, although differences between groups in these two CSE measures did not reach statistical significance. Sensitivity analyses revealed that reductions in active and resting excitability in the ipsilesional hemisphere were exacerbated in those individuals with subcortical lesions affecting the CST (**Supplementary Table 4**). Taken together, these findings align with previous studies showing that subcortical lesions lead to a greater overall reduction of excitability in the ipsilesional hemisphere.^14^ Furthermore, this reduction appears to be more substantial when descending motor pathways in subcortical areas are damaged.^28^

At face value, the lack of differences between groups in CSP, ICF and SICI observed in this study may seem at odds with a study of smaller size (n=40) but with a more precise characterization of the lesion location that showed both greater reductions in SICI (i.e. disinhibition) in individuals with cortical lesions, and longer prolongations of the CSP in individuals with subcortical lesions.^14^ That study, however, only included acute and very early subacute patients (<14 days), who typically exhibit greater reductions in SICI and increases in CSP than individuals in later stages of recovery.^29^ Our analyses included time since stroke as covariate to factorize the impact of stroke chronicity, which had little influence in the models (**Supplementary Tables 3and 5**). More importantly, comparisons between cortical and subcortical groups in the study mentioned previously^14^ were made using interhemispheric ratios (ipsilesional/contralesional) of these CSE measures and thus results cannot be directly compared with our study. In line with the findings of that study we found that, in individuals with cortical lesions, SICI was more reduced in the ipsilesional than in the contralesional hemisphere (**Table 2**). However, our results do not support the view that lesion location has a significant influence in GABA-mediated inhibition.^30^

We found striking differences in the associations between CSE measures obtained from the ipsilesional hemisphere and motor skill acquisition in individuals with cortical and subcortical lesions (**Fig. 4**). In individuals with cortical lesions, more excitability in the ipsilesional hemisphere, expressed as reduced RMT, was significantly associated with better motor skill acquisition. This association is consistent with the much-debated hypothesis that CSE levels predict motor performance and skill learning.^31^ In contrast, we found that in individuals with subcortical lesions, it was lower CSE expressed as smaller resting MEP amplitudes that was associated with better motor skill acquisition. This association was not expected because it is precisely in those individuals with more subcortical CST damage where smaller MEPs would be expected to be associated with poorer motor skill performance.^31^ The exact reason for this negative association is still to be elucidated but it could be related to the recruitment of alternative networks during motor skill practice.

Indeed, in individuals with subcortical lesions, motor skill learning has been associated with increased activity and connectivity in frontal brain regions alongside reduced activity in motor areas, implying a reliance on compensatory cortical changes to preserve learning capacity.^32^ It is therefore possible that, in these individuals, reduced CSE could reflect, paradoxically, an increased recruitment of alternative cortical circuits that support motor skill acquisition when CST connections are disrupted.^10^ Redistribution of synaptic strength and neural excitability within a dynamic range might be crucial for maintaining stability amongst motor networks.^33^ Subcortical lesions disrupting the CST can trigger increased activity in secondary motor regions and the recruitment of residual pathways as a compensatory mechanism to preserve corticospinal outputs.^10^

We also found marked differences between cortical and subcortical groups in the associations between motor skill acquisition and CSE measures of facilitation (ICF) and inhibition (SICI) in the ipsilesional hemisphere (**Fig. 4**). The primary motor cortex (M1) and its descending corticospinal projections are under GABAergic influence, with inhibitory neurons suppressing any excitatory drive emerging from the neighboring intracortical representations and thus regulating neuronal action potential firing.^34^ Following ischemia or hypoxic damage, phasic GABA-related signaling is dramatically reduced, unmasking normally suppressed connections and shifting the excitatory balance towards facilitation.^35^ Such changes in facilitatory-inhibitory balance have been shown to be critical for inducing neuroplastic structural changes that support recovery.^36^ In animal models with motor pathway lesions, reducing inhibition can lead to sustained motor recovery improvements.^37^ Multiple studies have attempted to investigate the functional implications of this excitatory-inhibitory balance in post-stroke survivors and the implication for motor recovery, but findings remain still inconclusive.^38^

Our study provides novel insights into the functional role of this excitatory-inhibitory balance by revealing the existence of two distinct intracortical excitability patterns underlying motor skill acquisition depending on lesion location. In individuals with cortical lesions, better acquisition rates were associated with greater inhibitory activity, suggesting that neurophysiological patterns similar to neurotypical individuals when lesions spare corticospinal pathways.^39^ Conversely, in subcortical lesions, where descending corticospinal fibers tend to be more severely compromised, a shift in the facilitation-inhibition balance might be needed in order to preserve motor skill acquisition capacity, with reduced intracortical inhibition unmasking latent excitatory glutamatergic connections.^40^ Our sensitivity analyses confirmed that both ICF and SICI associations in the subcortical group were primarily driven by damage in the CST because only in individuals with lesions in this pathway these associations remained significant (**Supplementary Table 8**). These findings reinforce previous evidence in support of the critical role that GABAergic and glutamatergic balance has in brain repair and recovery processes after stroke.^13^

Given the neuroanatomy of the CST, the fact that no statistically significant associations with motor skill acquisition were observed for any of the CSE measures obtained from the contralesional hemisphere was not unexpected (**Fig. 5**). This finding aligns with previous experimental and clinical studies indicating that improvements in upper-limb recovery derive from changes in activity taking place primarily in peri-infarct neurons in the ipsilesional hemisphere.^41^ Associations between CSE and the contralesional hemisphere may become significant only in individuals with more severe subcortical lesions who present a substantial or complete disruption of corticospinal connections.^15^ Since our study included mostly individuals with mild levels of upper-limb motor dysfunction (**Table 1**), a significant disruption of the CST projections possibly occurred only in a very small proportion of our participants.

The mechanistic insights discovered in this study could be relevant for clinical practice. Modifying CSE by either inhibiting or facilitating intracortical networks is the backbone of most non-invasive brain stimulation (NIBS) treatments aimed at improving upper-limb motor recovery after stroke. Despite multiple efforts, there is currently no clear evidence that these interventions benefit upper-limb recovery after stroke, in part because its treatment response can be influenced by numerous factors including lesion location.^42^ Consensus-based recommendations for NIBS in stroke rehabilitation highlight the need for increased understanding regarding response phenotypes and neural mechanisms in order to improve individualization of treatment protocols.^43^ Our results provide a neurophysiological rationale regarding NIBS-induced positive effects and perhaps assisting in identifying more accurate therapeutical targets to promote recovery.

## Limitations

The most important limitation of this study is the lack of structural neuroimaging data to better characterize the stroke lesion and the extend of the CST damage. To partly mitigate this limitation, we used exploratory sensitivity analyses to determine if having subcortical lesions affecting the CST impacted motor skill acquisition, CSE, and their associations. Obtaining data on the microstructural integrity (e.g., anisotropy) of the CST would have improved the interpretation of our results. Acknowledging the fact that brain lesions do not fit into a simple binary classification, our study allowed us to address a specific mechanistic question, demonstrating the important role of lesion location in mediating the neurophysiological properties underlying motor skill acquisition post-stroke.

A second limitation concerns the level of motor disability of the participants and how this limited the generalizability of our results to individuals with more severe upper-limb impairments. It is possible that people with more severe lesions could show different patterns in the associations between CSE and motor skill acquisition.^15^ Nevertheless, about 65% of stroke survivors exhibit coordination and control deficits in the affected hand, even in cases with mild or no motor impairment.^2^ Including participants with relatively moderate levels of upper-limb motor dysfunction allowed us to assess motor skill acquisition using a highly functional motor task and to obtain complete CSE measures from most participants. Obtaining TMS data in very impaired patients whose MEP responses from the ipsilesional hemisphere cannot be easily elicited is challenging.^44^ Future studies should validate to what extent our results can be extrapolated to more impaired individuals.

A third limitation refers to the lack of a delayed retention test of the time on target motor task to investigate the capacity to retain the gains in motor skill acquisition obtained during practice. Motor skill learning is a complex multistage process that requires encoding sensorimotor information during motor practice but also the consolidation of such information as procedural memory.^45^ To evaluate motor skill learning and distinguish it from potential transient improvements in skill performance occurring during motor practice a delayed retention test of the practiced motor task would be needed.^46^ Studies investigating whether associations between CSE measures and motor skill acquisition remain significant when retention is assessed and the influence of lesion location are warranted.^47^

## Conclusion

Understanding the basic neurophysiological mechanisms underlying distinct phases of motor skill learning after stroke has the potential to impact recovery and rehabilitation significantly.^16^ However, the ubiquitous heterogeneity in human stroke studies limits the identification of unique neural patterns that could improve the ability to predict long-term outcomes and responses to treatments.^48^ This study sheds light on this issue by showing the influential role of lesion location in upper-limb motor skill acquisition and its underlying neurophysiological mechanisms. Contrary to what we observed in motor impairment and function, motor skill acquisition was relatively well preserved in individuals with subcortical lesions, who, compared with individuals with cortical lesions, tended to show lower levels of excitability in the ipsilesional hemisphere. Furthermore, while better motor skill acquisition was associated with neurotypical excitatory patterns in cortical strokes, lower excitability, intracortical inhibition and higher facilitation correlated with larger acquisition rates in subcortical lesions. These results indicate that motor skill acquisition may rely on distinct corticospinal circuits depending on lesion location after stroke and suggest that alternative networks may play an important role in preserving acquisition in those lesions affecting descending corticospinal pathways. These findings offer valuable insights to identify more accurate therapeutical targets for the design of treatment approaches aimed at promoting recovery following stroke.

## Data Availability

Data is available upon reasonable request.

